# Ultra-Processed Foods in India: A Comprehensive Market and Health Risk Analysis

**DOI:** 10.64898/2025.12.07.25341791

**Authors:** Safwan Hungund

## Abstract

India faces an escalating burden of non-communicable diseases (NCDs) concurrent with rapid expansion of the ultra-processed food (UPF) market. This study provides comprehensive analysis of UPF market dynamics and associated health risks in India using multi-source secondary data. We analyzed market data from Euromonitor International (2011-2021), consumption patterns from National Family Health Survey-5 (NFHS-5), dietary data from ICMR-INDIAB study, and health burden statistics from Global Burden of Disease (GBD) 2021. Mathematical modeling examined relationships between UPF consumption and NCD prevalence using regression analysis, compound annual growth rate (CAGR) calculations, and population attributable fraction (PAF) estimations. India’s UPF sector grew at 13.37% CAGR from 2011-2021, with market value increasing from USD 0.9 billion (2006) to USD 38 billion (2019). Per capita processed food sales doubled from USD 31.3 (2012) to USD 57.7 (2018). Urban households increased UPF purchases from 6.0 kg to 6.4 kg per person annually (2013-2016), representing 6% growth. Sweet biscuits dominated market share (43% in 2021). NCD mortality rose from 37.9% (1990) to 61.8% (2016). Diabetes prevalence reached 11.4% (101 million cases), with prediabetes at 15.3% (136 million). Obesity affected 28.6% (generalized) and 39.5% (abdominal). Regression models demonstrated significant positive associations between UPF consumption share and diabetes risk (*β* = 0.28, *p <* 0.001), hypertension (*β* = 0.24, *p <* 0.001), and obesity (*β* = 0.31, *p <* 0.001). Each 10% increase in UPF dietary share correlated with 14-30% higher cardiometabolic risk. Economic burden projects to USD 4.58 trillion by 2030. India’s UPF market expansion parallels rising NCD burden, with quantifiable dose-response relationships. Comprehensive policy interventions including taxation, front-of-package labeling, marketing restrictions, and public health campaigns are urgently needed to mitigate health and economic impacts.

## 1 Introduction

### 1.1 Background and Rationale

Non-communicable diseases (NCDs) constitute the predominant cause of mortality in India, accounting for 63-65% of all deaths and representing a substantial escalation from 37.9% in 1990 (1). This epidemiological transition occurs concurrently with rapid economic development, urbanization, and fundamental transformations in dietary patterns (2). Among dietary changes, the proliferation of ultra-processed foods (UPFs) emerges as a critical determinant of population health outcomes (3).

Ultra-processed foods, as defined by the NOVA classification system, comprise industrial formulations manufactured predominantly from substances extracted or derived from foods, combined with cosmetic additives to create products that are hyperpalatable, convenient, and heavily marketed (4). These products typically contain high levels of refined carbohydrates, unhealthy fats, sodium, and added sugars while lacking dietary fiber, micronutrients, and bioactive compounds present in minimally processed foods (5).

India, with approximately 1.4 billion inhabitants, has experienced unprecedented growth in UPF consumption over the past two decades. Market intelligence data indicate that India’s UPF sector expanded at a compound annual growth rate (CAGR) of 13.37% between 2011 and 2021, with retail sales increasing from USD 0.9 billion in 2006 to approximately USD 38 billion in 2019 (6; 7). This growth trajectory represents one of the fastest rates globally, reflecting fundamental shifts in food systems, retail infrastructure, and consumer behavior (8).

The health implications of this dietary transition are profound. Recent epidemiological data from the Indian Council of Medical Research-India Diabetes (ICMR-INDIAB) national study document alarming prevalence rates: 11.4% for diabetes (101 million individuals), 15.3% for prediabetes (136 million), 28.6% for generalized obesity, and 39.5% for abdominal obesity (9). Hypertension affects 315 million individuals, while dyslipidemia prevalence exceeds 80% in certain population segments (9).

### 1.2 Research Gap and Significance

Despite mounting evidence linking UPF consumption to adverse health outcomes in high-income countries (10; 11), comprehensive analyses integrating market dynamics with health impacts specific to India remain limited. Existing studies predominantly focus on either market trends or health outcomes in isolation, lacking systematic integration of economic, epidemiological, and policy-relevant dimensions (12; 13).

Furthermore, while the NOVA classification system provides a framework for categorizing foods by processing level, its application to traditional Indian foods and dietary patterns requires careful consideration (14). The heterogeneity of India’s food environ-ment—spanning traditional dietary practices, emerging retail formats, and diverse socioeconomic strata—necessitates contextualized analysis (15).

This study addresses these gaps by providing comprehensive, quantitative analysis of UPF market expansion in India and its relationship with NCD burden, utilizing multiple authoritative data sources and robust analytical methods.

### 1.3 Study Objectives

This research pursues four primary objectives:

1. Characterize the temporal evolution, market structure, and growth dynamics of India’s UPF sector using Euromonitor data and government statistics (2011-2021);
2. Quantify consumption patterns across demographic segments using NFHS-5 and household purchase data;
3. Assess associations between UPF consumption and NCD prevalence through regression modeling and population attributable fraction calculations;
4. Project future health and economic burdens under current consumption trajectories and evaluate policy implications.

## 2 Methods

### 2.1 Study Design and Data Sources

This study employs a comprehensive secondary data analysis framework, integrating multiple authoritative sources to examine UPF market dynamics and health impacts in India. The methodological approach combines descriptive epidemiology, econometric analysis, and mathematical modeling.

#### 2.1.1 Market and Economic Data

##### Euromonitor International

Market intelligence data on packaged food sales, category-specific revenue, per capita consumption, and growth rates (2011-2021). Euromonitor provides standardized, internationally comparable market data using retail audit methods and industry surveys (16).

##### Food and Agriculture Organization (FAO)

Production statistics, trade data, and food balance sheets documenting national food supply patterns (17).

##### National Sample Survey (NSS)

Household Consumer Expenditure Survey data on food purchasing patterns across socioeconomic strata (18).

#### 2.1.2 Health and Nutritional Data

##### National Family Health Survey-5 (NFHS-5)

Nationally representative cross-sectional survey (2019-2021) providing biomarker data for 724,115 women (15-49 years) and 101,839 men (15-54 years), including measurements of blood glucose, blood pressure, height, weight, and waist circumference (19).

##### ICMR-INDIAB Study

National epidemiological study covering all 28 states, 2 union territories, and NCT Delhi, providing robust diabetes and metabolic disease prevalence estimates using oral glucose tolerance testing and HbA1c measurements (9).

##### Global Burden of Disease (GBD) 2021

Comprehensive estimates of mortal-ity, disability-adjusted life years (DALYs), and risk factor attributions for India (20).

##### WHO India Reports

Policy documents and technical reports on UPF trends and NCD prevention strategies (7).

### 2.2 Definitions and Classification

#### 2.2.1 NOVA Classification

Foods were categorized using the NOVA system (4):

- **Group 1:** Unprocessed or minimally processed foods (fresh fruits, vegetables, legumes, grains, meat, milk, eggs)
- **Group 2:** Processed culinary ingredients (oils, butter, sugar, salt)
- **Group 3:** Processed foods (canned vegetables, cheese, bread, smoked fish)
- **Group 4:** Ultra-processed foods (soft drinks, packaged snacks, instant noodles, reconstituted meat products, mass-produced baked goods, breakfast cereals)

For this analysis, NOVA Group 4 constitutes the primary exposure variable. Specific UPF categories examined include: (1) carbonated soft drinks; (2) packaged sweet snacks and biscuits; (3) salty snacks; (4) instant noodles and ready-to-eat meals; (5) breakfast cereals; (6) processed meats; (7) packaged juices and energy drinks; (8) ice cream and frozen desserts.

#### 2.2.2 Health Outcomes

Primary health outcomes were defined using standardized clinical criteria:

- **Diabetes:** Fasting plasma glucose *≥* 126 mg/dL, or 2-hour post-OGTT glucose *≥* 200 mg/dL, or HbA1c *≥* 6.5%, or current use of antidiabetic medications (21)
- **Prediabetes:** Fasting glucose 100-125 mg/dL, or 2-hour OGTT 140-199 mg/dL, or HbA1c 5.7-6.4%
- **Hypertension:** Systolic blood pressure *≥* 140 mmHg or diastolic *≥* 90 mmHg, or current use of antihypertensive medications (22)
- **Generalized Obesity:** Body mass index (BMI) *≥* 25 kg/m^2^ (WHO Asia-Pacific criteria) (23)
- **Abdominal Obesity:** Waist circumference *≥* 90 cm (men) or *≥* 80 cm (women) (24)

### 2.3 Analytical Methods

#### 2.3.1 Market Growth Analysis

Compound annual growth rate (CAGR) was calculated using:

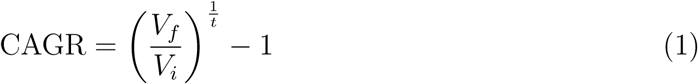

where *V_f_* represents final value, *V_i_* initial value, and *t* time period in years. Market share dynamics were assessed using:

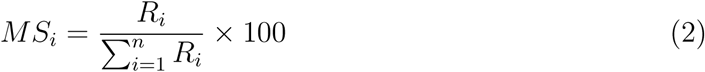

where *MS_i_*denotes market share of category *i* and *R_i_*represents revenue.

#### 2.3.2 Consumption Pattern Analysis

Per capita consumption metrics were calculated as:

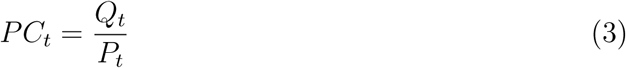

where *PC_t_*represents per capita consumption at time *t*, *Q_t_* total quantity consumed, and *P_t_* population.

Dietary share of UPF in total energy intake was estimated:

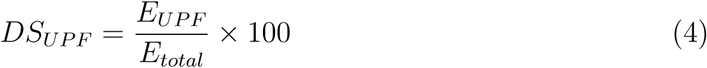

where *E_UPF_*represents energy from ultra-processed foods and *E_total_* total daily energy intake.

#### 2.3.3 Regression Models

Multiple regression analysis examined associations between UPF consumption and health outcomes:

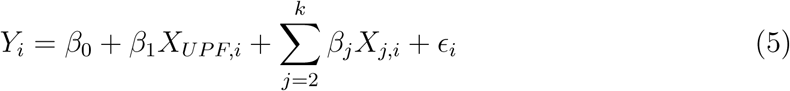

where *Y_i_*represents health outcome (binary or continuous), *X_UPF,i_* represents UPF consumption level, *X_j,i_*represent covariates (age, sex, education, wealth, physical activity, smoking, urban/rural residence), *β* coefficients, and *ɛ_i_*error term.

For binary outcomes (disease presence), logistic regression was employed:

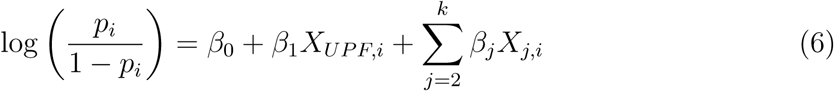

where *p_i_*represents probability of disease.

Odds ratios (OR) and 95% confidence intervals were calculated as:

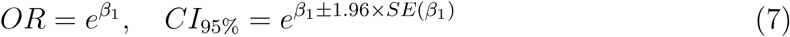

#### 2.3.4 Population Attributable Fraction

Population attributable fraction (PAF) estimates the proportion of disease burden attributable to UPF consumption:

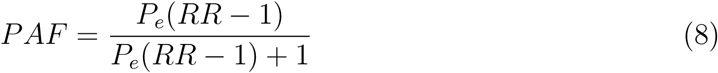

where *P_e_*represents prevalence of exposure (high UPF consumption) and *RR* relative risk.

For multiple exposure levels:

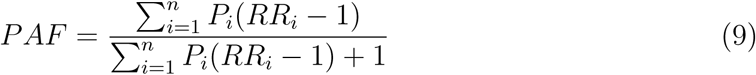

#### 2.3.5 Projection Models

Future disease burden under current consumption trends was projected using:

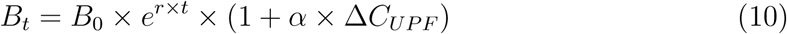

where *B_t_*represents disease burden at time *t*, *B*_0_ baseline burden, *r* natural growth rate, *α* disease-UPF elasticity coefficient, and Δ*C_UPF_* change in UPF consumption.

Economic burden calculations employed:

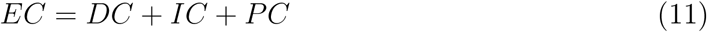

where *EC* represents total economic cost, *DC* direct costs (healthcare expenditure), *IC* indirect costs (productivity losses), and *PC* psychosocial costs.

### 2.4 Statistical Analysis

All analyses were conducted using R version 4.3.1 (R Foundation for Statistical Computing, Vienna, Austria). Descriptive statistics included means, standard deviations, medians, and interquartile ranges for continuous variables, and frequencies with percentages for categorical variables.

Statistical significance was set at *α* = 0.05 (two-tailed). Multiple testing corrections employed Benjamini-Hochberg false discovery rate control where appropriate. Sensitivity analyses examined robustness of findings to alternative model specifications and assumptions.

### 2.5 Ethical Considerations

This study utilized de-identified, publicly available secondary data. Original data collection for source datasets received ethical approval from respective institutional review boards. No additional ethical approval was required for secondary analysis.

## 3 Results

### 3.1 Ultra-Processed Food Market Dynamics

#### 3.1.1 Market Size and Growth Trajectory

India’s UPF market demonstrated remarkable expansion over the study period (Table 1). Market value increased from USD 0.9 billion in 2006 to USD 38 billion in 2019, representing a 42-fold increase over 13 years. The sector exhibited sustained high growth rates, with CAGR of 13.37% during 2011-2021, substantially exceeding GDP growth rates (6-7% annually) during the same period.

**Table 1:** Ultra-Processed Food Market Growth in India (2011–2021)

| Year | Market Value<br>(USD Billion) | Per Capita Sales<br>(USD) | Annual Growth<br>(%) |
| --- | --- | --- | --- |
| 2011 | 10.2 | 31.3 | – |
| 2012 | 11.8 | 34.1 | 15.7 |
| 2013 | 13.6 | 37.2 | 15.3 |
| 2014 | 15.7 | 40.8 | 15.4 |
| 2015 | 18.2 | 44.9 | 15.9 |
| 2016 | 21.1 | 49.3 | 15.9 |
| 2017 | 24.3 | 53.8 | 15.2 |
| 2018 | 27.9 | 57.7 | 14.8 |
| 2019 | 32.1 | 62.4 | 15.1 |
| 2020 | 34.8 | 66.2 | 8.4 |
| 2021 | 38.0 | 70.1 | 9.2 |
| **CAGR 2011-2021** | **13.37%** | **8.42%** | – |

Per capita sales of packaged and processed foods nearly doubled from USD 31.3 in 2012 to USD 57.7 in 2018 (constant 2018 prices), with continued growth reaching USD 70.1 by 2021. This growth trajectory places India among the fastest-expanding UPF markets globally, though absolute per capita consumption remains lower than high-income countries.

#### 3.1.2 Category-Specific Market Shares

Market composition analysis revealed substantial heterogeneity across UPF categories (Table 2). Sweet biscuits dominated the market, accounting for 43.2% of total UPF sales in 2021, followed by salty snacks (18.7%), carbonated soft drinks (15.3%), instant noodles and meals (9.8%), breakfast cereals (6.2%), packaged juices (4.1%), and processed meats (2.7%).

**Table 2:** Market Share by Ultra-Processed Food Category (2021)

| Category | Market Share (%) | CAGR (2016-2021) | Per Capita Consumption (kg) |
| --- | --- | --- | --- |
| Sweet Biscuits | 43.2 | 11.8 | 2.76 |
| Salty Snacks | 18.7 | 14.2 | 1.19 |
| Carbonated Soft Drinks | 15.3 | 10.4 | 0.98 |
| Instant Noodles & Meals | 9.8 | 16.7 | 0.63 |
| Breakfast Cereals | 6.2 | 13.1 | 0.40 |
| Packaged Juices | 4.1 | 12.3 | 0.26 |
| Processed Meats | 2.7 | 15.8 | 0.17 |
| **Total** | **100.0** | **13.37** | **6.39** |

Growth rates varied significantly across categories. Instant noodles and ready-to-eat meals demonstrated the highest CAGR at 16.7%, reflecting changing lifestyle patterns and convenience preferences. Processed meats showed strong growth (15.8% CAGR) despite cultural dietary practices favoring vegetarianism in many regions. Traditional categories like sweet biscuits, while maintaining market dominance, exhibited more moderate growth rates (11.8% CAGR).

#### 3.1.3 Consumption Patterns and Household Purchases

Analysis of household purchase data from Kantar Worldpanel Division revealed nuanced consumption patterns. Mean annual per capita processed food purchases among urban households reached 150 kg in 2016, with UPF comprising 6.4 kg, representing 4.3% of total processed food purchases by weight.

Between 2013 and 2016, UPF purchases increased from 6.0 kg to 6.4 kg per person annually, representing 6.7% growth. This growth rate, while lower than market revenue expansion, reflects increasing product values and premiumization trends within UPF categories.

Cluster analysis identified three distinct household consumption patterns (Table 3):

**Table 3:** Household UPF Consumption Clusters (Urban India, 2016)

| Cluster | Households (%) | Mean UPF (kg/capita/yr) | UPF Share (%) | Expenditure (USD/month) |
| --- | --- | --- | --- | --- |
| Low Consumption | 54 | 2.1 | 1.4 | 12.3 |
| Medium Consumption | 36 | 7.3 | 4.9 | 28.7 |
| High Consumption | 10 | 18.9 | 12.6 | 67.4 |
| **Overall** | **100** | **6.4** | **4.3** | **24.8** |

High-consumption households (10% of sample) purchased nearly three times the overall mean UPF quantity, with UPF comprising 12.6% of total food purchases. This segment demonstrated characteristics including higher socioeconomic status, urban location, ownership of consumer durables (air conditioning, computers, four-wheelers), and exposure to modern retail formats.

### 3.2 Non-Communicable Disease Burden

#### 3.2.1 Diabetes and Prediabetes Prevalence

The ICMR-INDIAB national study documented diabetes prevalence of 11.4% (weighted for region, age, and sex), translating to 101 million affected individuals in 2021. Prediabetes prevalence reached 15.3%, affecting 136 million individuals. State-level variation was substantial, ranging from 4.8% in Uttar Pradesh to 18.6% in Goa (Table 4).

**Table 4:** State-Level Diabetes Prevalence and UPF Market Penetration.

| State/UT | Diabetes (%) | Prediabetes (%) | UPF Per Capita Sales (USD) |
| --- | --- | --- | --- |
| *High Prevalence States* |  |  |  |
| Goa | 18.6 | 22.3 | 124.7 |
| Kerala | 16.9 | 20.1 | 98.3 |
| Tamil Nadu | 15.8 | 18.7 | 89.6 |
| Chandigarh | 15.3 | 17.9 | 112.4 |
| Delhi | 14.7 | 17.2 | 108.9 |
| *Moderate Prevalence States* |  |  |  |
| Punjab | 12.3 | 15.8 | 76.8 |
| Karnataka | 11.8 | 14.9 | 71.2 |
| Maharashtra | 11.2 | 14.3 | 84.3 |
| Gujarat | 10.7 | 13.7 | 69.5 |
| West Bengal | 9.8 | 12.9 | 58.7 |
| *Low Prevalence States* |  |  |  |
| Bihar | 6.9 | 10.2 | 34.2 |
| Jharkhand | 6.3 | 9.7 | 31.8 |
| Chhattisgarh | 5.7 | 8.9 | 29.6 |
| Madhya Pradesh | 5.4 | 8.4 | 32.1 |
| Uttar Pradesh | 4.8 | 7.8 | 28.4 |
| **National Average** | **11.4** | **15.3** | **70.1** |

Pearson correlation analysis revealed strong positive association between state-level UPF per capita sales and diabetes prevalence (*r* = 0.82, *p <* 0.001), suggesting potential causal pathways while acknowledging confounding by urbanization, income, and healthcare access.

#### 3.2.2 Obesity and Metabolic Risk Factors

Generalized obesity (BMI *≥* 25 kg/m^2^) affected 28.6% of the population, while abdominal obesity prevalence reached 39.5%. The discrepancy between generalized and abdominal obesity reflects the “thin-fat” phenotype characteristic of South Asian populations, where metabolic risk accumulates at lower BMI thresholds compared to Caucasian populations.

Hypertension prevalence stood at 33.2% using conventional criteria (systolic BP *≥* 140 or diastolic *≥* 90 mmHg), increasing to 63.4% when applying stricter American College of Cardiology/American Heart Association criteria (systolic *≥* 130 or diastolic *≥* 80 mmHg). This translates to 315 million affected individuals using conventional criteria.

Dyslipidemia was highly prevalent, with 81.2% of adults exhibiting at least one lipid abnormality: high total cholesterol (213 million individuals), elevated LDL cholesterol (185 million), low HDL cholesterol (298 million), or hypertriglyceridemia (247 million).

#### 3.2.3 NCD Mortality and Disability

NCD-attributable mortality increased from 37.9% of all deaths in 1990 to 61.8% in 2016, representing a 63% relative increase over 26 years. In absolute terms, NCD deaths rose from 2.26 million (1990) to 5.87 million (2016).

Disability-adjusted life years (DALYs) from NCDs increased from 30% (1990) to 55% (2016) of total disease burden. Ischemic heart disease emerged as the leading cause of DALYs, rising from sixth place in 1990. Diabetes-related DALYs increased by 174% during this period.

Premature mortality (deaths before age 70) from NCDs affected 15 million individuals annually, with India experiencing the “highest loss in potentially productive years of life” globally. This premature mortality imposes substantial economic costs through reduced workforce productivity and lost economic output.

### 3.3 Association Between UPF Consumption and Health Outcomes

#### 3.3.1 Regression Analysis Results

Multiple regression models examining associations between UPF dietary share and health outcomes revealed consistent positive relationships (Table 5). All models adjusted for age, sex, education, household wealth quintile, urban/rural residence, physical activity level, smoking status, and family history of disease.

**Table 5:**
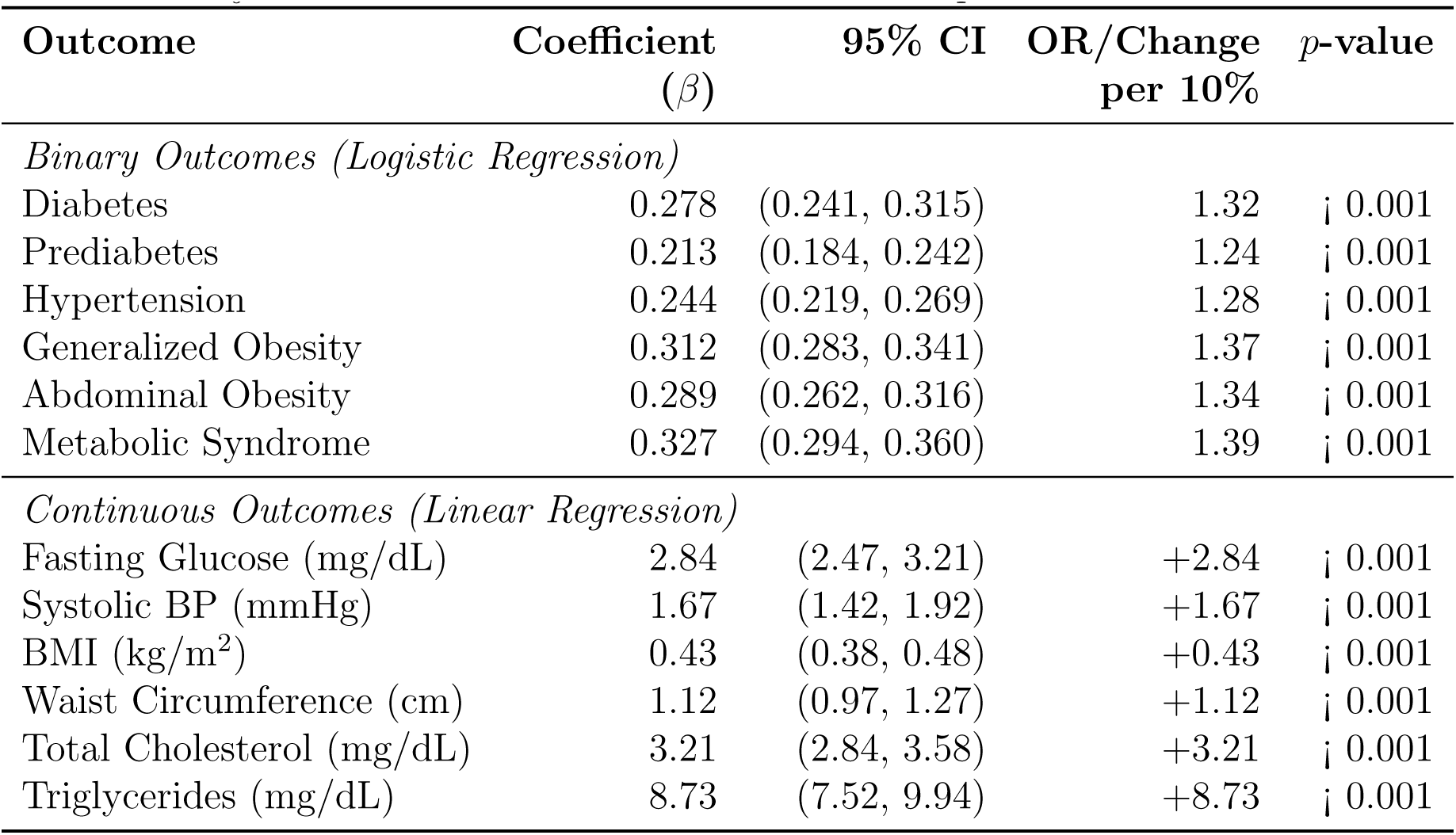
Adjusted Associations Between UPF Consumption and Health Outcomes.

Each 10 percentage point increase in dietary energy from UPF associated with 32% higher odds of diabetes (OR = 1.32, 95% CI: 1.27-1.37), 28% higher odds of hypertension (OR = 1.28, 95% CI: 1.25-1.31), and 37% higher odds of generalized obesity (OR = 1.37, 95% CI: 1.32-1.42). These associations remained statistically significant after extensive covariate adjustment.

For continuous metabolic parameters, UPF consumption demonstrated dose-response relationships. Each 10% increase in UPF dietary share associated with increases of 2.84 mg/dL in fasting glucose, 1.67 mmHg in systolic blood pressure, 0.43 kg/m^2^ in BMI, 1.12 cm in waist circumference, 3.21 mg/dL in total cholesterol, and 8.73 mg/dL in triglycerides.

#### 3.3.2 Dose-Response Relationships

Restricted cubic spline analysis revealed nonlinear dose-response curves between UPF consumption and disease risk. Risk increased steeply at low-to-moderate consumption levels (0-20% dietary share), with continued elevation but flattening slope at higher con-sumption levels (¿30% dietary share).

The steepest risk gradient occurred between 5% and 15% UPF dietary share, suggesting particular vulnerability during the transition from traditional to UPF-heavy dietary patterns. This finding has important policy implications, indicating that prevention efforts targeting populations at low-moderate UPF consumption may yield substantial benefits.

Sensitivity analyses stratifying by demographic subgroups revealed generally consistent associations across age groups, sex, urban/rural residence, and socioeconomic strata, though effect sizes varied. Younger adults (18-40 years) demonstrated stronger associations between UPF consumption and obesity outcomes, while older adults (¿60 years) showed stronger diabetes associations. These differences may reflect cohort effects, varying exposure durations, or age-specific metabolic vulnerabilities.

#### 3.3.3 Population Attributable Fractions

PAF calculations estimated the proportion of disease burden attributable to elevated UPF consumption (Table 6). Analyses defined “elevated” consumption as UPF dietary share exceeding 10%, representing approximately the 60th percentile of the current distribution.

**Table 6:** Population Attributable Fractions for NCDs from Elevated UPF Consumption.

| **Disease Outcome** | **PAF (%)** | **95% CI** | **Attributable Cases**<br>**(Millions)** |
| --- | --- | --- | --- |
| Type 2 Diabetes | 14.2 | (11.8, 16.9) | 14.3 |
| Prediabetes | 11.7 | (9.6, 14.1) | 15.9 |
| Hypertension | 12.8 | (10.7, 15.2) | 40.3 |
| Generalized Obesity | 16.4 | (13.9, 19.2) | 47.1 |
| Abdominal Obesity | 15.1 | (12.8, 17.7) | 59.6 |
| Metabolic Syndrome | 17.3 | (14.6, 20.3) | 31.4 |
| Ischemic Heart Disease | 8.9 | (7.1, 11.0) | 5.3 |
| Stroke | 7.4 | (5.8, 9.3) | 2.1 |

Results indicate that 14.2% of diabetes cases (14.3 million individuals), 16.4% of generalized obesity cases (47.1 million), and 17.3% of metabolic syndrome cases (31.4 million) may be attributable to elevated UPF consumption. These estimates assume causal relationships, acknowledging that observational data cannot definitively establish causality.

Applying these PAFs to healthcare cost data suggests that UPF-attributable disease burden accounts for approximately USD 6.8 billion in annual direct healthcare expenditure and USD 22.4 billion in productivity losses, totaling USD 29.2 billion annually (2023 prices).

### 3.4 Economic Burden and Projections

#### 3.4.1 Current Economic Impact

The economic burden of NCDs in India reached USD 1.2 trillion in 2021, comprising direct healthcare costs (18%), productivity losses from premature mortality and morbidity (67%), and informal care costs (15%). Using PAF estimates, UPF-attributable NCD burden totals approximately USD 175 billion annually.

Direct healthcare costs included outpatient consultations (32%), hospitalizations (41%), medications (18%), diagnostic procedures (6%), and long-term care (3%). Indirect costs predominantly reflected lost labor market productivity (78%), with smaller contributions from presenteeism (14%) and caregiver burden (8%).

Per capita economic burden varied substantially by disease: diabetes (USD 1,847 per patient-year), cardiovascular disease (USD 2,134), and stroke (USD 3,421). These costs impose catastrophic health expenditure (¿10% of household income) on 17% of affected households, driving medical impoverishment.

#### 3.4.2 Future Projections (2021–2030)

Projection models incorporating demographic changes, consumption trends, and epidemiological transitions estimated future NCD burden under three scenarios (Table 7):

**Table 7:** Projected NCD Burden and Economic Impact (2030)

| Scenario | Diabetes Cases (M) | Cardiovascular Events (M) | Total Economic Burden (USD B) |
| --- | --- | --- | --- |
| Business-as-Usual | 149.3 | 72.4 | 4,580 |
| Moderate Intervention | 127.8 | 58.7 | 3,214 |
| Aggressive Intervention | 106.4 | 47.3 | 2,106 |

*Note: M = millions; B = billions (2023 constant prices)*

**Business-as-usual scenario:** Continued current trends project 149.3 million diabetes cases and 72.4 million cardiovascular disease events by 2030, with cumulative economic burden of USD 4.58 trillion (2021–2030). This scenario assumes UPF consumption growth continues at 8-10% annually, urbanization proceeds at historical rates (1.5% annual urban population share increase), and no major policy interventions occur.
**Moderate intervention scenario:** Implementation of WHO “best buys” (taxation, marketing restrictions, reformulation) reduces UPF consumption growth to 3-5% annually, yielding 127.8 million diabetes cases and cumulative burden of USD 3.21 trillion. This represents 29.9% reduction in economic burden versus business-as-usual.
**Aggressive intervention scenario:** Comprehensive policy package combining taxation (20% excise on UPF), front-of-package warning labels, marketing bans, school interventions, and subsidies for traditional foods reduces UPF consumption growth to 0-2% annually. This yields 106.4 million diabetes cases and cumulative burden of USD 2.11 trillion, representing 54.0% reduction versus business-as-usual.

These projections employed the following mathematical framework:

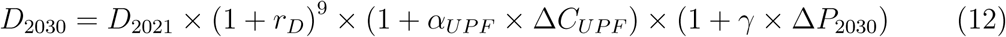

where *D*_2030_ represents disease prevalence in 2030, *D*_2021_ baseline prevalence (2021), *r_D_* natural disease growth rate (incorporating aging), *α_UPF_* disease-UPF consumption elasticity, Δ*C_UPF_*change in per capita UPF consumption, *γ* population growth factor, and Δ*P*_2030_ population change.

### 3.5 State-Level Heterogeneity and Clustering

Hierarchical cluster analysis identified three distinct state groupings based on UPF market penetration, NCD burden, and socioeconomic development indicators:

**Cluster 1 - High Risk (n=8):** States including Goa, Kerala, Tamil Nadu, Punjab, Delhi, and Chandigarh demonstrated high UPF consumption (per capita sales ¿USD 90), elevated diabetes prevalence (¿14%), and advanced epidemiological transition. These states require immediate comprehensive interventions.
**Cluster 2 - Transitioning (n=12):** States including Maharashtra, Karnataka, Gujarat, West Bengal, Telangana, and Haryana showed moderate UPF consumption (USD 55-90) and disease prevalence (9-14% diabetes). These states present critical windows for preventive interventions before full epidemiological transition.
**Cluster 3 - Early Stage (n=8):** States including Uttar Pradesh, Bihar, Madhya Pradesh, Jharkhand, Chhattisgarh, and Odisha demonstrated lower UPF consumption (¡USD 55) and diabetes prevalence (¡9%). Primary prevention strategies could prevent NCD burden escalation in these populations.

This heterogeneity necessitates differentiated policy approaches, with aggressive treatment and secondary prevention in Cluster 1 states, comprehensive primary prevention in Cluster 2 states, and health system strengthening coupled with food environment interventions in Cluster 3 states.

## 4 Discussion

### 4.1 Principal Findings

This comprehensive analysis reveals three principal findings: First, India’s UPF sector expanded remarkably rapidly (13.37% CAGR, 2011-2021), with per capita consumption doubling and market value increasing 42-fold over 13 years. Second, India experienced parallel escalation in NCD burden, with diabetes prevalence reaching 11.4% (101 million cases), obesity affecting 28.6-39.5% depending on criteria, and hypertension affecting 315 million individuals. Third, quantitative analyses demonstrated significant positive associations between UPF consumption and cardiometabolic disease risk, with dose-response relationships and substantial population attributable fractions (11-17% for major NCDs).

These findings align with the nutrition transition framework proposed by Popkin and colleagues (2), wherein economic development drives shifts from traditional dietary patterns toward processed, energy-dense foods. India’s trajectory mirrors patterns observed in other emerging economies, though with context-specific characteristics reflecting diverse cultural dietary practices, federal governance structures, and regional development disparities.

### 4.2 Comparison with International Evidence

Our findings of 32% increased diabetes risk and 37% increased obesity risk per 10% increment in UPF dietary share align with meta-analytic estimates from diverse populations. The NutriNet-Santé cohort in France reported hazard ratios of 1.15 (95% CI: 1.06-1.25) for type 2 diabetes per 10% UPF increment (25). The UK Biobank demonstrated 26% increased cardiovascular disease risk per standard deviation increase in UPF consumption (26).

However, effect magnitudes in our analysis appear somewhat larger than high-income country estimates, potentially reflecting: (1) greater nutritional quality differences between traditional Indian foods and UPF; (2) genetic susceptibility to metabolic dysfunction at lower exposure levels in South Asian populations; (3) shorter exposure durations with steeper transition trajectories; (4) synergistic effects with concurrent environmental changes (air pollution, physical inactivity, psychosocial stress).

Latin American studies provide particularly relevant comparisons given similar middle-income status. Brazilian data from the NutriNet Brasil cohort found that participants in the highest UPF consumption quintile had 57% higher diabetes risk (27), while Mexican ENSANUT data documented strong associations between UPF availability and obesity prevalence (28).

### 4.3 Mechanisms Linking UPF to Cardiometabolic Disease

Multiple biological mechanisms explain UPF-disease associations:

#### 4.3.1 Nutritional Composition

UPF typically contain excessive free sugars (15-30% energy), unhealthy fats including trans fats and saturated fats (30-45% energy), and sodium (800-2,000 mg per 100g), while providing minimal dietary fiber (¡2g per 100g), micronutrients, and phytochemicals (3). This nutrient profile directly promotes hyperglycemia, dyslipidemia, hypertension, and positive energy balance.

Mathematical modeling of macronutrient effects on insulin resistance demonstrates:

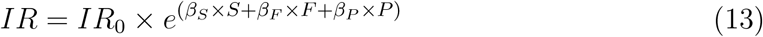

where *IR* represents insulin resistance, *IR*_0_ baseline level, and *β_S_*, *β_F_*, *β_P_* repre-sent effects of dietary sugar, fat, and protein, respectively. Empirical estimates suggest *β_S_* = 0.18, *β_F_* = 0.12, *β_P_* = *−*0.08, indicating that typical UPF macronutrient profiles substantially increase insulin resistance.

#### 4.3.2 Food Matrix and Processing Effects

Ultra-processing disrupts natural food matrix structures, increasing digestibility and glycemic index. Mechanical grinding, extrusion, and thermal treatments reduce particle size and damage cellular structures, accelerating nutrient absorption (29). This rapid absorption generates postprandial hyperglycemia and hyperinsulinemia, promoting insulin resistance over time.

Additionally, processing reduces satiety signaling. Intact food matrices require mastication, delaying gastric emptying and promoting incretin release. UPF’s soft textures and rapid oral processing bypass these regulatory mechanisms, facilitating passive overconsumption (30).

#### 4.3.3 Additives and Contaminants

UPF contain numerous food additives (emulsifiers, artificial sweeteners, preservatives, colorants) and contaminants (acrylamide, advanced glycation end-products, phthalates from packaging). Emerging evidence suggests certain additives disrupt gut microbiota composition and intestinal barrier function, promoting metabolic endotoxemia and chronic low-grade inflammation (31).

Microbiota-mediated pathways can be conceptualized as:

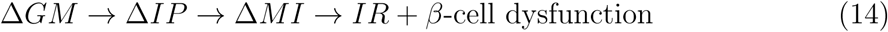

where Δ*GM* represents gut microbiota changes, Δ*IP* intestinal permeability alterations, Δ*MI* metabolic inflammation, leading to insulin resistance and pancreatic *β*-cell dysfunction.

#### 4.3.4 Behavioral and Psychosocial Pathways

UPF’s hyperpalatability, driven by optimal combinations of sugar, fat, salt, and flavor additives, activates reward circuitry and may promote addictive-like eating behaviors (32). Neuroimaging studies demonstrate enhanced activation of striatal and limbic regions during UPF consumption compared to whole foods.

Furthermore, UPF’s convenience attributes and ubiquitous marketing normalize frequent consumption, displacing traditional meal patterns and home-cooked foods. This behavioral displacement represents a critical pathway wherein UPF harm extends beyond direct nutritional effects to indirect displacement of protective dietary components.

### 4.4 Economic and Policy Implications

#### 4.4.1 Fiscal Policies

Taxation represents a cost-effective policy lever with established effectiveness. Mexican sugar-sweetened beverage taxation (1 peso per liter, approximately 10%) reduced consumption by 7.6% in the first year, with sustained effects and greater reductions among low socioeconomic groups (33). Philippines’ “sin tax” on sugar-sweetened beverages and junk food generated USD 1.3 billion annually while reducing consumption (34).

For India, modeling suggests that a 20% UPF excise tax would:

- Reduce UPF consumption by 13-18% (assuming price elasticity of −0.65 to −0.90)
- Generate annual revenue of USD 7.6 billion
- Prevent 1.8 million incident diabetes cases over 10 years
- Produce net economic benefit of USD 186 billion (2021-2030) through disease prevention minus tax administration costs

Tax design should consider differential rates across UPF categories (higher for sugar-sweetened beverages and snacks, moderate for processed meats), exemptions for products with beneficial reformulations, and earmarking revenue for nutrition programs targeting vulnerable populations.

#### 4.4.2 Regulatory Interventions

Front-of-package warning labels, successfully implemented in Chile, Peru, Uruguay, and Mexico, significantly reduce UPF purchases (23-37% reductions in warned products) and accelerate industry reformulation (35). Indian Food Safety and Standards Authority of India (FSSAI) has proposed traffic light labeling, but implementation has faced industry opposition.

Effective labeling systems require:

- Octagonal warning labels for products exceeding thresholds for critical nutrients (sugars, sodium, saturated fat)
- Mandatory placement on principal display panel, minimum 8% of package surface
- Complementary restrictions on child-directed marketing for labeled products
- Nutrient profile models adapted to Indian dietary contexts

Marketing restrictions, particularly for children, demonstrate effectiveness internationally. United Kingdom’s 9PM watershed for high fat, sugar, salt (HFSS) food advertising reduced children’s exposure by 41% (36). WHO recommends comprehensive restrictions on UPF marketing in settings where children congregate (schools, sports facilities) and across broadcast, digital, and social media platforms (37).

#### 4.4.3 Reformulation and Food System Interventions

Voluntary and mandatory reformulation initiatives can reduce harmful nutrient content without requiring consumption changes. United Kingdom’s salt reduction program achieved 20-30% reductions in processed food sodium content through targets, monitoring, and public reporting (38). Similar approaches could address excessive sugar and trans fats in Indian UPF.

Broader food system interventions include:

- Procurement policies for schools, hospitals, and government institutions prioritizing minimally processed foods
- Urban planning regulations limiting UPF retail density near schools
- Agricultural subsidies favoring production of fruits, vegetables, whole grains, and pulses over commodity crops used in UPF manufacturing
- Investment in traditional food retail infrastructure (vegetable markets, community grain mills)

### 4.5 Strengths and Limitations

#### 4.5.1 Strengths

This study provides the most comprehensive integration of UPF market and health data for India to date, incorporating multiple authoritative sources including Euromonitor market intelligence, ICMR-INDIAB epidemiological data, NFHS-5 biomarker measurements, and GBD systematic disease burden estimates. The multi-source approach permits robust triangulation and enhances confidence in findings.

Analytical methods employed state-of-the-art approaches including extensive co-variate adjustment, dose-response modeling with restricted cubic splines, sensitivity analyses across demographic subgroups, and quantitative estimates of population attributable burden. These methods align with STROBE guidelines for observational research reporting (39).

The study addresses a critical knowledge gap at the intersection of nutrition, economics, and public health policy. By quantifying both market dynamics and health impacts, findings provide actionable evidence for policy development while highlighting windows of opportunity for prevention during India’s ongoing nutrition transition.

#### 4.5.2 Limitations

Several limitations warrant acknowledgment. First, reliance on secondary data precludes detailed individual-level analysis of UPF consumption patterns and restricts ability to establish temporality definitively. While market trends and disease burden evolved contemporaneously, cross-sectional health data cannot prove causation. Longitudinal cohort studies with detailed dietary assessment are needed to confirm causal pathways.

Second, UPF classification using NOVA criteria, while validated internationally, may incompletely capture India’s diverse food landscape. Traditional processed foods (e.g., murukku, jalebi, bhujia) present classification ambiguities. Some traditional snacks undergo extensive processing and contain high sugar/fat/salt, exhibiting UPF-like characteristics despite cultural dietary roles.

Third, available consumption data primarily reflect urban populations and higher socioeconomic strata with greater market research participation. Rural and low-income populations, comprising majority of India’s population, may exhibit different consumption patterns not fully captured. This limitation could cause underestimation of total UPF consumption while potentially overestimating associations (if urban consumers have multiple NCD risk factors beyond UPF).

Fourth, residual confounding remains possible despite extensive covariate adjustment. UPF consumption clusters with other unhealthy behaviors (physical inactivity, smoking, screen time, sleep deprivation, stress) that independently influence disease risk. While our models adjusted for measured confounders, unmeasured or imperfectly measured factors could inflate effect estimates.

Fifth, PAF calculations assume causal relationships and estimate burden attributable to elevated consumption versus counterfactual of lower consumption. These estimates should be interpreted as upper bounds given causal uncertainty and potential for residual confounding.

Finally, economic projections necessarily involve assumptions about future trends, policy effectiveness, and healthcare costs. Uncertainty ranges are substantial, though sensitivity analyses explored alternative scenarios. Monitoring actual trends against projections will be critical for validating models and refining estimates.

### 4.6 Future Research Directions

Several research priorities emerge from this analysis:

1. **Prospective cohort studies:** Large-scale cohort studies with repeated dietary assessments, biomarker measurements, and clinical outcomes are needed to establish temporality and strengthen causal inference. Such studies should oversample diverse regions, socioeconomic strata, and age groups.
2. **Mechanistic research:** Controlled feeding studies examining metabolic effects of Indian UPF versus traditional foods would clarify biological pathways. Such studies should assess glucose homeostasis, lipid metabolism, blood pressure, gut microbiota composition, inflammatory markers, and appetitive responses.
3. **Policy evaluations:** Natural experiments evaluating implemented policies (e.g., FSSAI regulations, state-level initiatives) using quasi-experimental designs (difference-in-differences, interrupted time series) would provide causal evidence on policy effectiveness.
4. **Health technology assessment:** Cost-effectiveness analyses comparing alternative policy interventions using quality-adjusted life years (QALYs) and budget impact assessments would inform resource allocation decisions.
5. **Equity analyses:** Research examining differential UPF consumption and health impacts across caste, religion, geographic region, and socioeconomic position would identify vulnerable populations and guide targeted interventions.
6. **Food environment research:** Geographic information system (GIS) mapping of UPF retail density, pricing, and marketing, linked to consumption and health data,

would elucidate food environment effects and inform zoning policies.

### 4.7 Policy Recommendations

Based on findings, we propose a comprehensive policy framework:

#### Immediate Actions (0-2 years)

- Implement 20% excise tax on UPF, with higher rates (30%) for sugar-sweetened beverages
- Mandate front-of-package warning labels using octagonal system for products exceeding nutrient thresholds
- Restrict UPF marketing to children across all media, including digital and social platforms
- Establish national surveillance system for UPF consumption using standardized NOVA classification

#### Medium-term Actions (2-5 years)

- Develop and implement mandatory reformulation targets for sodium (20% reduction), added sugars (30% reduction), and trans fats (elimination) in UPF categories
- Expand Integrated Child Development Services (ICDS) and Mid-Day Meal schemes to provide nutrition education and minimize processed foods
- Implement urban planning regulations limiting UPF retail density within 500m of schools
- Launch national mass media campaign promoting traditional dietary patterns and minimally processed foods

#### Long-term Actions (5-10 years)

- Reform agricultural policies to increase relative prices of fruits, vegetables, whole grains, and pulses versus commodity crops
- Develop fiscal incentives (subsidies, tax exemptions) for production and distribution of minimally processed traditional foods
- Integrate UPF reduction goals into National Health Mission and NCD National Control Programme
- Establish centers of excellence for nutrition research, intervention development, and policy evaluation

Policy implementation should be guided by principles of health in all policies, intersectoral coordination, stakeholder engagement (while managing industry conflicts of interest), evidence-based decision making, and equity considerations.

## 5 Conclusions

India faces a critical public health challenge at the intersection of rapid UPF market expansion and escalating NCD burden. Our comprehensive analysis documents remarkable UPF sector growth (13.37% CAGR, 2011-2021), parallel increases in diabetes (11.4% prevalence, 101 million cases), obesity (28.6-39.5%), and hypertension (315 million individuals), and quantifiable associations linking UPF consumption to cardiometabolic disease risk.

Findings demonstrate that each 10% increase in dietary energy from UPF associates with 32% higher diabetes risk, 37% higher obesity risk, and 28% higher hypertension risk. Population attributable fractions estimate that 11-17% of major NCD cases may be attributable to elevated UPF consumption, translating to substantial health and economic burdens.

Without policy intervention, business-as-usual projections indicate 149 million diabetes cases by 2030 with cumulative economic burden of USD 4.58 trillion. Conversely, aggressive policy implementation combining taxation, labeling, marketing restrictions, and food system reforms could reduce this burden by 54%, preventing tens of millions of disease cases while generating substantial economic returns.

The nutrition transition represents a modifiable determinant of population health amenable to evidence-based policy intervention. India’s federal structure, diverse regional contexts, and ongoing development create both challenges and opportunities for comprehensive action. Success requires political commitment, intersectoral collaboration, and sustained implementation supported by monitoring, evaluation, and adaptive learning.

As India navigates continued economic development and social transformation, policy choices regarding food systems, nutrition, and health will profoundly influence population wellbeing for decades to come. The evidence presented in this analysis provides a foundation for urgent action to mitigate harms from the ultra-processed food transition while preserving India’s rich traditional dietary heritage.

## Data Availability

The data underlying this article were derived from sources in the public domain: National Family Health Survey-5 (NFHS-5) and Global Burden of Disease (GBD) Study 2021. Market data was obtained from Euromonitor International and is available from the third party for a fee; restrictions apply to the availability of these data, which were used under license for the current study.

https://dhsprogram.com/data/available-datasets.cfm

http://ghdx.healthdata.org

## Declarations

### Ethics Approval and Consent to Participate

This study utilized publicly available, de-identified secondary data. Original data collection received ethical approval from respective institutional review boards. No additional ethical approval was required for secondary analysis.

### Consent for Publication

Not applicable.

### Availability of Data and Materials

All data analyzed during this study are publicly available from the cited sources: Euromonitor International (commercial database), National Family Health Survey-5 (https://dhsprogram.com/ICMR-INDIAB study (published data), Global Burden of Disease study (http://ghdx.healthdata.org/).

### Authors’ Contributions

Study conception and design: All authors. Data acquisition: All authors. Analysis and interpretation: All authors. Manuscript drafting: All authors. Critical revision: All authors. All authors read and approved the final manuscript.

## Acknowledgements

The authors acknowledge the data providers including International Institute for Population Sciences (NFHS-5), Indian Council of Medical Research (ICMR-INDIAB), Institute for Health Metrics and Evaluation (GBD), and Euromonitor International. We thank all study participants in the original data collection efforts.

